# Altered Neurotransmitter Ratio in the Prefrontal Cortex is Associated with Pain in Fibromyalgia Syndrome

**DOI:** 10.1101/2021.10.28.21265618

**Authors:** James H. Bishop, Afik Faerman, Andrew Geoly, Naushaba Khan, Claudia Tischler, Heer Amin, Adi Maron-Katz, Azeezat Azeez, David C. Yeomans, Ralph Hurd, Meng Gu, Laima Baltusis, Daniel Spielman, Matthew D. Sacchet, David Spiegel, Nolan Williams

## Abstract

The central mechanisms underlying fibromyalgia syndrome (FMS) remain undetermined. The dorsolateral prefrontal cortex (DLPFC) is particularly relevant to FMS because it is implicated in cognitive, affective, and top-down pain regulation. Imbalances in excitatory (Glutamate) and inhibitory (Gamma aminobutyric acid; GABA) neurochemicals may play a critical role in the pathophysiology of the condition and more generally in homeostatic function within cortical circuits. Although the balance of excitation and inhibition are intrinsically linked no investigations to date have investigated the E/I ratio in FMS. Thus, the primary objective of this study was to determine whether the E/I ratio in the DLPFC is altered in participants with FMS compared to healthy controls using magnetic resonance spectroscopy. Additionally, we examined the relationship between E/I ratio and pain metrics. We hypothesized that the E/I ratio within the DLPFC would be altered in participants with FMS compared to controls and, secondly, that E/I ratio would be associated with both clinical pain and thermal pain sensitivity. The Brief Pain Inventory (BPI) self-assessment was used to evaluate pain severity and impact on physical functioning and acute pain sensitivity was determined via quantitative sensory testing to define thermal (heat) pain threshold and tolerance. Our results revealed an elevation in the E/I ratio in FMS compared to controls. A positive relationship between E/I ratio and thermal pain sensitivity measures was identified in the FMS cohort. Collapsing across groups, there was a positive relationship between E/I ratio and BPI score. These findings suggest that dysfunction in the balance between excitation and inhibition within cognitive brain circuitry may play a role in pain processing in FMS.

## INTRODUCTION

Fibromyalgia syndrome (FMS) is a poorly understood disorder of unknown etiology hallmarked by widespread pain, fatigue, and sleep disturbances, cognitive and emotional dysregulation, as well as increased mechanical and thermal sensitivity [4; 8; 80]. The mechanisms underlying the condition remain unclear, however, increasing evidence suggests that neurochemical imbalances factor into the pathophysiology of the disorder [18; 31; 63]. Magnetic resonance spectroscopy (MRS) enables non-invasive quantification of multiple metabolites that are present in the brain in millimolar concentrations (0.5-10mM) [76; 84]. Among the different neurochemicals that can be evaluated by MRS, Glx, a combination of glutamate and glutamine, and gamma-aminobutyric acid (GABA) are considered to be the primary excitatory and inhibitory neurotransmitters respectively [60]. Alterations in both Glx and GABA within limbic brain regions have been demonstrated in FMS and across chronic pain disorders (see reviews; [44; 57]) including elevated Glx and reduced GABA in the insular cortex [18; 31], a structure involved in pain intensity coding and modulation [71]. Complementary molecular imaging approaches (e.g., positron emission tomography; PET) have provided additional evidence of cortical GABA alterations in FMS indicated by increased GABAA receptor concentrations [8]. Collectively, these studies indicate dysregulation in both excitatory and inhibitory systems in FMS which are intrinsically linked to one another by the glutamate/GABA-glutamine cycle [3], however, this clinically relevant synergistic relationship has yet to be characterized.

In addition to the gap described above, the majority of MRS studies in FMS examine primary sensory and affective circuitry necessitating additional investigations in higher order cortical structures known to be involved in chronic pain conditions. Specifically, the dorsolateral prefrontal cortex (DLPFC) plays a pivotal role in mediating cognitive, evaluative, and sensory components of the pain experience and is involved in top-down pain modulation [68] via cortico-subcortical and cortico-cortical pathways [47]. For example, transcranial magnetic stimulation (TMS) of both the right and left DLPFC (R/L-DLPFC) have been indicated to provide FMS symptom relief [42; 69; 72; 82]. Although the mechanisms underlying TMS effects are debated, inhibitory forms of stimulation have been shown to increase GABA [70]. Characterizing the balance of excitation to inhibition within the DLPFC is an important step toward understanding the pathophysiology of the condition.

Thus, in this investigation, we examined whether baseline DLPFC alterations in the ratio of excitatory (Glx) to inhibitory (GABA) neurochemical concentrations (E/I ratio) were present between FMS and controls using single proton MRS (^1^H-MRS). Left DLPFC (L-DLPFC) MRS was collected and analyzed. Given that FMS is frequently comorbid with other medical and psychiatric disorders [22; 33; 45], we conducted an exploratory analysis dichotomizing the FMS cohort by subjective report of depression in the past 6-months. As secondary analyses, we examined the relationship between the E/I ratio, clinical pain scores, and acute thermal (heat) pain sensitivity. Consistent with previous reports highlighted above demonstrating increased Glx and reduced GABA across several brain regions in FMS, we predicted that E/I ratio would be elevated in the L-DLPFC compared to controls. Based on findings of elevated Glx and reduced GABA, we hypothesized that the E/I ratio would be associated with clinical pain and acute pain sensitivity measured by a thermal pain threshold and tolerance assay. Although many studies investigating FMS either report solely on female populations or exclude males because of the skewed prevalence across sex [20], this sentiment has recently been challenged [81], and thus we conducted two independent analyses: 1) males and females combined and 2) females only (due to lack of sufficient sample size for male only comparison). Identifying whether combined neurochemical imbalances are present in centralized pain disorders such as FMS is pertinent to identify potential mechanisms of novel therapies particularly given the limited treatment options for the condition.

## METHODS

### Participant Recruitment and Inclusion

Participants with FMS and controls were recruited separately as part of two independent studies. Both studies were approved by the Stanford University Institutional Review Board (IRB). Prior to enrollment, all FMS participants underwent phone and in-person screening procedures. Participants with chronic pain were evaluated by a study physician to confirm a primary clinical diagnosis of FMS. Diagnostic criteria were determined based on the American College of Rheumatology Preliminary Diagnostic Criteria for FMS and additionally participants were required to have blood samples within the last two months confirming normal complete blood count (CBC) and inflammatory panel. Exclusionary criteria for the FMS cohort include MRI contraindications, neurological disorders (i.e., seizure disorder, insomnia, etc.), and psychological disorders (i.e., bipolar disorder, schizophrenia, etc.). Given the high comorbidity rate of depression and chronic pain syndromes, particularly FMS [45], depression was not deemed exclusionary, however participants with unmanaged depressive disorder or depression with suicidal ideation were excluded from the study. Because data for this study was collected in conjunction to an ancillary transcranial magnetic stimulation (TMS) trial for FMS, participants taking psychoactive medications such as antidepressants and/or opioid analgesics were required to undergo a washout period to mitigate seizure risks. Washout was done prior to imaging and was individually tailored to the participant by the study clinician. Healthy controls inclusion and exclusion criteria were identical to the FMS cohort, however, individuals with a past or present history of chronic pain were excluded. Prior to imaging, participants underwent urinalysis drug and pregnancy screening and if positive were excluded from further procedures.

### Study Design

MRS data in the FMS cohort was collected supplementary to an ongoing clinical trial. Following confirmed eligibility and enrollment, participants with FMS underwent a baseline MRI session (MRI #1) to identify a personalized L-DLPFC functional connectivity cluster similar to our previous report [11]. Each participant’s individualized cluster represented the area within the L-DLPFC exhibiting the greatest functional connectivity to the dorsal anterior cingulate cortex (dACC). The cluster was then used to guide subsequent brain stimulation (not included in this manuscript) as well as the placement of the MRS voxel in the FMS cohort (Figure 1). FMS participants then underwent two separate experimental days each consisting of two independent MRI sessions sandwiched between the application of brain stimulation (MRI #’s 2-5). Only pre-TMS MRS acquisitions were analyzed and reported in this manuscript. Control participants underwent a single baseline MRI session (MRI #1) where the L-DLPFC MRS voxel was prescribed at the center-of-gravity anatomical coordinate of the BA9+BA46 mask (Figure 1). See *Magnetic Resonance Imaging Data Acquisition* section for more details. A total of 81 participants (FMS = 59; Controls = 22) had MRS scans with 13 excluded due to low data quality (shim error / fit error >12% / linewidth >12Hz; FMS = 7; Control = 6) and scanning related issues (i.e., falling asleep during scans; FMS =1), yielding a total analyzable sample of 67 (FMS = 51; Controls = 16) (Table 1; Figure 1; Supplementary Figure 1 - Consort Diagram). Due to the differing experimental designs of the studies, there was an opportunity to repeat poor quality MRS scans in the FMS group at the second follow-up visit (if warranted) resulting in less data attrition than controls.

**Table 1.** Demographics, Behavioral/Clinical Variables, and MRS Data Quality.

| **Demographics & Behavioral / Clinical Variables** |  |  |  |
| --- | --- | --- | --- |
| (N = 67; Fibromyalgia = 51, Controls = 16) |  |  |  |
|  | **Fibromyalgia** | **Control** | **P value** |
| Age (Years) | 48.73 (13.15) | 38.56 (13.01) | 0.009* |
| Sex (Females/Males) | 46 / 5 | 12 / 4 | 0.12 |
| Current Pain (Day of Experiment; 0-10) | 4.31 (2.05) | 0 (0) | 4.616<sup>-09*</sup> |
| Average Pain (Day of Experiment; 0-10) | 4.53 (1.92) | 0 (0) | 4.841<sup>-09*</sup> |
| Brief Pain Inventory (BPI) | 19.06 (6.24) | 3.00 (3.97) | 5.867<sup>-09*</sup> |
| Acute Thermal (Heat) Pain Threshold (°C) | 44.61 (3.55) | - | - |
| Acute Thermal (Heat) Pain Tolerance (°C) | 47.75 (2.51) | - | - |

| **MRS Data Quality** |  |  |  |
| --- | --- | --- | --- |
| (*3 FMS participants missing linewidth data) |  |  |  |
|  | **Fibromyalgia** | **Control** | **P value** |
| Linewidth (Hz) | 8.89 (1.80) | 8.25 (0.68) | 0.26 |
| Fit Error GABA / Cr (%) | 7.26 (1.51) | 8.27 (1.81) | 0.055 |
| Fit Error GABA / Water (%) | 7.09 (1.55) | 8.13 (1.84) | 0.052 |
| Fit Error Glx / Cr (%) | 6.19 (1.60) | 6.99 (1.73) | 0.12 |
| Fit Error Glx / Water (%) | 6.00 (1.64) | 6.84 (1.77) | 0.1 |

*Values for each variable except sex are presented as mean and standard deviation.*

**Figure 1.**
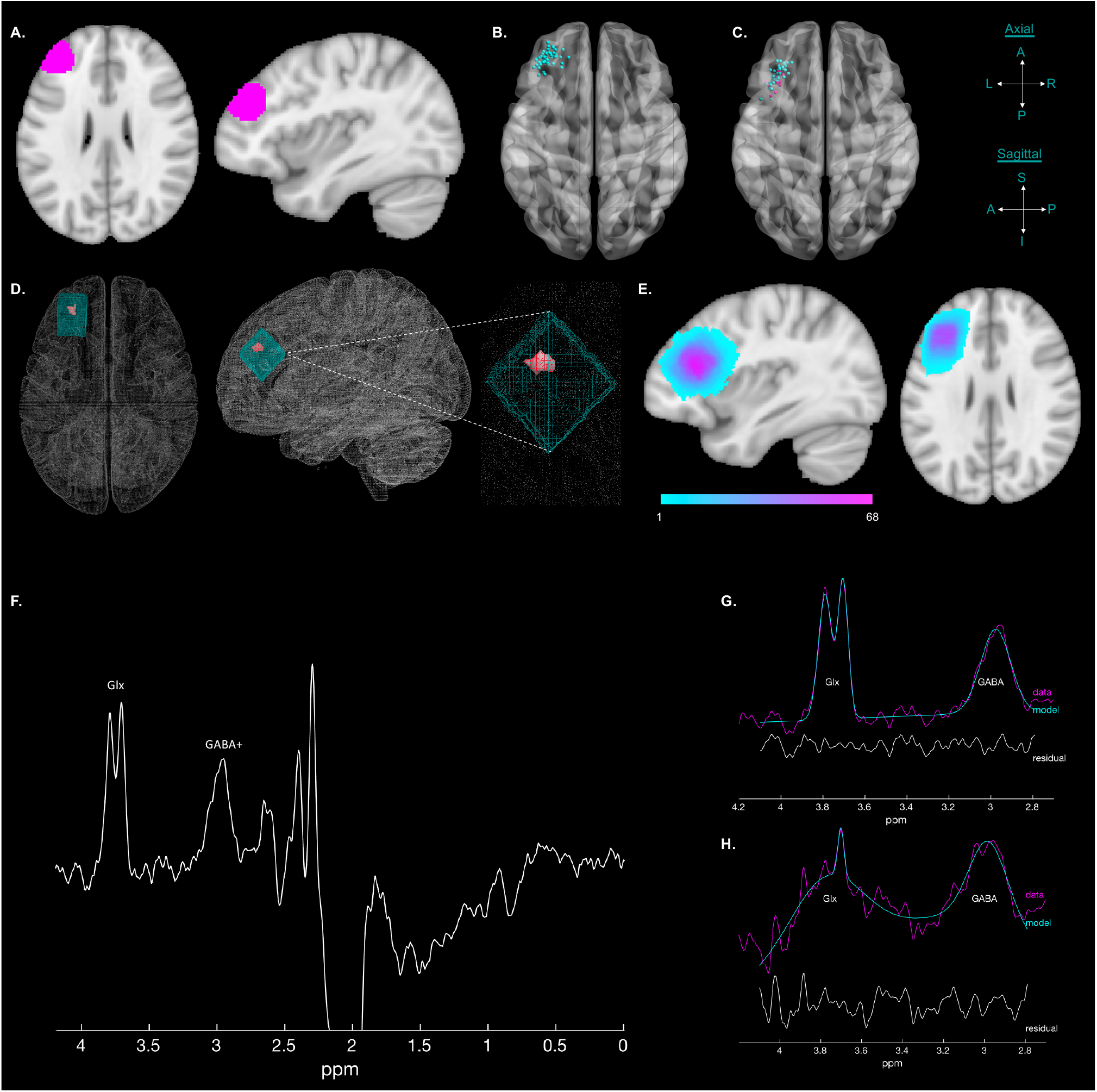
Schematic of L-DLPFC MRS Voxel Placement and Representative Data: A) Axial and sagittal L-DLPC region of interest for functional connectivity analyses consisting of BA 46 and BA 9. B) Center of gravity coordinate for each participant with FMS indicated the greatest resting L-DLPFC-dACC resting state functional connectivity which was used downstream for MRS voxel placement. C) Since control participants did not undergo multiple MRI sessions to enable resting state guided voxel prescription, the center of the combined BA 46 + BA 9 mask was used. The figure depicts the adjusted center of gravity voxel coordinates for the acquired 2cm^3^ MEGA-PRESS MRS in participants with FMS (cyan) and controls (pink). Fine adjustments were made prior to data collection to ensure the MRS voxel consisted solely of brain tissue - grey matter, white matter, and cerebral spinal fluid (CSF). D) Representative subject illustrating the MEGA-PRESS voxel and identified L-DLPFC-dACC fMRI cluster overlap. E) Heatmap of all participants (FMS and controls) voxels depicted on a standard brain template demonstrating spatial consistency. F) Representative edited MEGA-PRESS spectrum. G) Example of a processed MEGA-PRESS spectrum with “Good Fit” or <12% fit error. H) Example of a processed MEGA-PRESS spectrum with “Bad Fit” or >12% fit error.

### Acute Thermal Pain Sensitivity Testing and Brief Pain Inventory Self-Report

FMS participants underwent quantitative sensory testing during a designated study visit outside of the MRI environment. Acute thermal pain sensitivity was evaluated using an ascending method of limits protocol by applying a computer-controlled Peltier probe (30×30mm) to the left forearm with an advanced thermal stimulator (Medoc Pathway Model ATS). Each thermal stimulus gradually increased in temperature beginning at 32°C and ramping at a rate of 1°C/second. A safety cut-off limit was set at 52°C. Pain threshold (the onset of pain) and pain tolerance (highest level of pain the participant could tolerate) were determined. Patients were blinded from observing the temperatures to ensure an objective measure of pain sensitivity. The thermode was shifted between three discrete positions on the left forearm, within the same dermatome to avoid sensitization and habituation. Participants indicated their response by clicking a mouse button held in their contralateral (right) hand which initiated the machine to stop ramping and return to baseline. This task was repeated five times for pain threshold testing and three times for pain tolerance testing. Pain threshold and tolerances temperatures were then determined by calculating the average of the five and three trials respectively (Table 1).

Clinical pain was assessed in both control and FMS cohorts by self-report using the Brief Pain Inventory short form (BPI-sf) questionnaire [9]. The survey was administered during the screening evaluation prior to experimental procedures. Pain severity score was determined by averaging questions 3-6 which describes current pain as well as average pain, least pain, and worst pain in the last 24 hours. Each question consists of an 11-point scale with 0 indicating no pain to 10 indicating the worst pain imaginable (Table 1).

### Magnetic Resonance Imaging Data Acquisition

MRI data was obtained at the Center for Neurobiological Imaging at Stanford University using a research dedicated 3.0T General Electric Discovery MR750 instrument with a Nova Medical 32-channel head coil. Structural and metabolic acquisition sequences and parameters were identical for both FMS and control cohorts.

MRI sequence parameters were: 1) Whole-brain structural imaging consisted of a 0.9mm^3^ three-dimensional T1-weighted MPRAGE sequence; 2) A 2D MEscher-Garwood Point REsolved Spectroscopy (MEGA-PRESS; [50]) sequence with voxel size=20×20×20mm^3^ (volume=8mL); echo time (TE) 68 ms; repetition time (TR) 2 s; Larmor frequency = 1.76 MHz; spectral width (SW) = 5000 Hz; spectral points = 4096; number of excitations (NEX) = 128; number of averages (NA) = 256; editing pulses pulses applied at 1.9 ppm (ON) and 7.46 ppm (OFF); FOV = 24mm; slice thickness = 20mm; slice spacing = 40mm; automatic 1^st^ order shimming along x, y, and z gradients for a total acquisition time of 9m12s. Motion was mitigated by head-immobilization cushioning. 3) Whole-brain fMRI (resting state) was collected using a simultaneous multi-slice EPI sequence with the following parameters: TE=30ms, TR=2000ms, Flip angle=77°, slice acceleration factor=3, matrix=128×128, 1.8×1.8mm^2^ in-plane resolution, slice thickness=1.8mm, FOV=230×230mm, 87 contiguous axial slices.

### Identification of L-DLPFC Subunit for Guided MRS Voxel Prescription (FMS Only)

Participants with FMS were part of large clinical trial involving the identification of individualized fMRI targets for subsequent transcranial magnetic stimulation intervention similar to the method described by Cole and colleagues [11]. For this manuscript all imaging data was collected prior to any application of brain stimulation. Given the repeated acquisition paradigm of the fibromyalgia study, a voxelwise analysis of the rs-fMRI scan was analyzed from a previous imaging session to determine the subregion of the L-DLPFC (Broadman Area 9 + 46) exhibiting the greatest functional connectivity to the dorsal anterior cingulate (dACC). The identified functional cluster was subsequently utilized to guide MRS voxel placement for each participant using a custom semi-automated prescription pipeline (described in detail in the voxel prescription procedures section; Figure 1) [5]. Control participants underwent a single neuroimaging session and thus the L-DLPFC voxel prescription was guided using a similar semi-automated approach, instead using the center-of-gravity coordinate of the combined Brodmann Area 9 and 46 ROI mask that was used to constrain the functional analyses described in the fibromyalgia study (Figure 1).

### Voxel Prescription Procedures

MRS voxel prescription was done using an in-house real-time semi-automated voxel prescription pipeline that enabled alignment of the MRS voxel based on identified center-of-gravity functional or anatomical cartesian coordinates while the participant is in the scanner using spmcoregister (SPM12 Software; [2]) [5]. In the FMS cohort, immediately following completion of the T1-weighted image, a custom script was used to pull the data from the imaging server and reconstruct the T1-weighted scan to nifti format. The baseline (MRI #1) T1-weighted image was coregistered to the pre-TMS (MRI #2) T1-weighted image and the subsequent warp was applied to the coordinate representing the center-of-gravity (mm) of the resting state defined L-DLPFC ROI. Resting state preprocessing and identification of the L-DLPFC subunit was performed similarly to our previous reports [11]. Voxel rotation was aligned to the slope of the skull in the sagittal plane and adjusted inferiorly, if necessary, to ensure the prescription was constrained entirely within the brain and did not include skull or meninges. This process was identical for control participants except the baseline T1-weighted image was coregistered to the Montreal Neurological Institute (MNI) standard brain template and the geometric center of the combined left hemisphere Brodmann Area 9 and 46 mask (MNI Standard Space Coordinates: X=-36.7, Y=41.5, Z=25.8).

### MEGA-PRESS Preprocessing

MEGA-PRESS data was analyzed using Gannet software (Version 3.0) implemented within MATLAB (Figure 1). GABA+ and Glx analysis processing steps include: 1) processing of raw time-domain data from the scanner into GABA-edited frequency-domain difference (DIFF) spectra with the following steps – line broadening, zerofill, fast Fourier transform, and extraction of water frequency; 2) Spectra fitting to quantify the edited GABA signals. To perform GABA J-difference editing two acquisitions were acquired differing in the manipulation of the GABA spin system. Subsequent subtraction of the manipulated acquisitions revealed GABA signals under the more prominent creatine (Cr) signal at 3 ppm. Difference editing was applied by subtracting the larger Cr signal to identify the smaller GABA signal. Next, Cr signal frequency-domain fitting occurred in the editing-OFF spectra. This was conducted because the small GABA signal is susceptible to magnet instabilities including scanner drift and experimental quandaries such as subject movement. Quantification of GABA spectra were obtained by applying a Gaussian model with linear baseline to the GABA signals in the DIFF spectra constrained within frequencies from 2.8 to 3.6 ppm, a Lorentzian model for the Cr signals in the OFF spectra constrained between 2.7 and 3.1 ppm, and a Guassian-Lorentzian model for the unsuppressed water spectrum constrained between 2.8 and 3.6 ppm. The resulting outputs include the integral ratio of GABA+ relative to Cr, GABA+ concentration relative to water in institutional units (i.u.).

The resulting spectra were individually quality controlled. Spectra with a fit error, representing the standard deviation of the residuals expressed as a percentage of the signal amplitude, >12% were discarded from subsequent analyses (Table 1 & Figure 1) [13; 64]. Excitatory / inhibitory ratio is reported as log value of Glx/GABA referenced to both Cr and water. To account for between subject variability in metabolite concentrations attributable to differences in brain tissue composition within the MRS voxels (Figure 2), tissue corrected E/I ratios were derived in Gannet Software using the segmented T1-weighted image.

**Figure 2.**
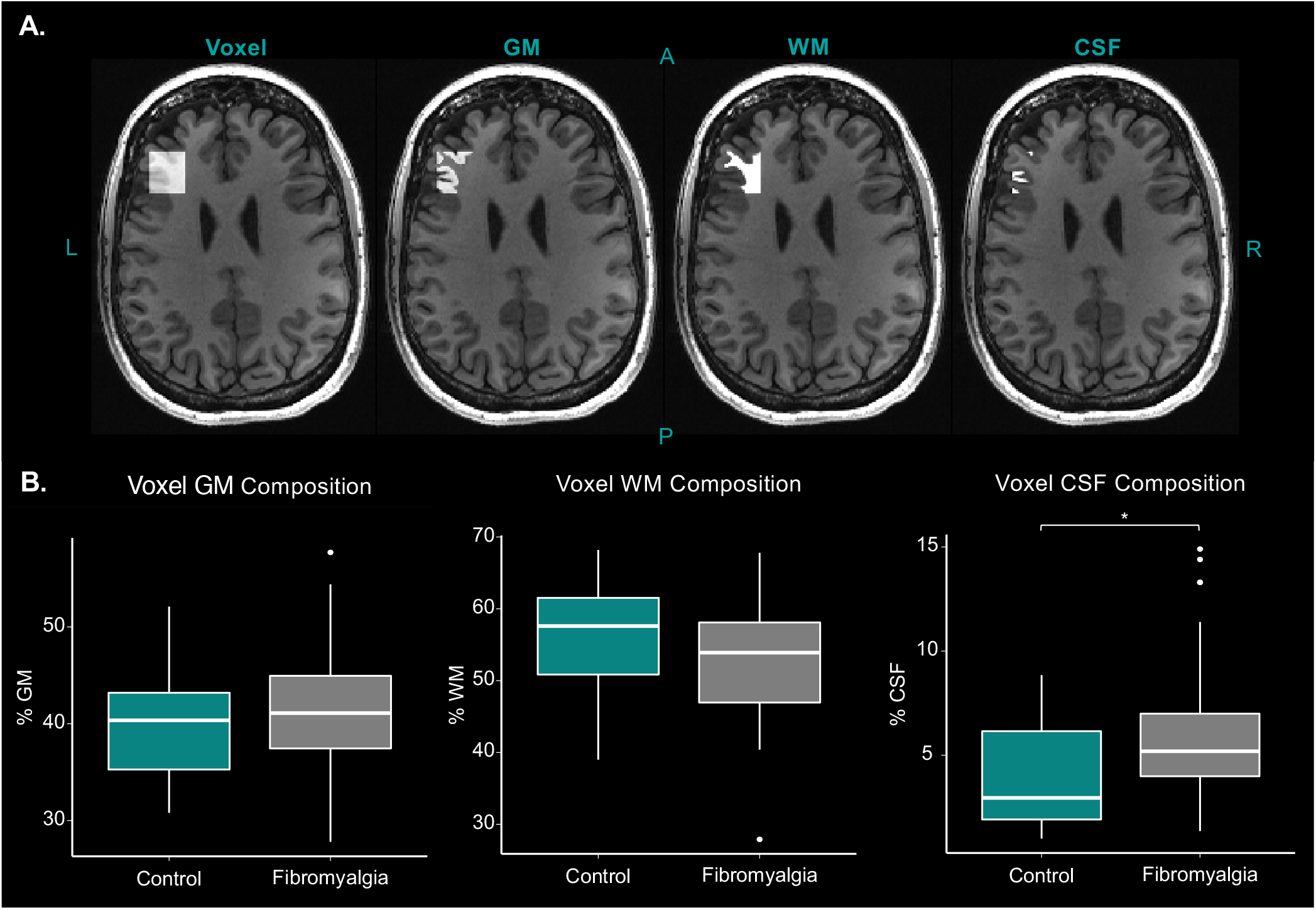
Voxel Composition: Neurochemical concentrations can be influenced by brain tissue composition within the MRS voxel. Segmentation of the voxel of interest was performed for all participants and used to correct across the sample as well as to assess potential differences between control and pain cohorts. A) Segmentations were conducted using Freesurfer software to determine the percentage of each tissue type with a representative single subject example shown above. B) Between group comparisons of brain tissue type indicating no differences between either white or grey matter, however, there was a significant between group difference in percentage of CSF.

### Statistical Analyses

T-tests were used to compare baseline mean E/I ratios between participants with FMS and controls. Multiple linear regression models were used to test the relationships between thermal pain sensitivity (tolerance and threshold) and E/I ratios in reference to both water and creatine (Cr) using IBM SPSS Statistics for Windows, version 26 (IBM Corp., Armonk, N.Y., USA). Pearson correlations were conducted to determine the relationship between L-DLPFC E/I ratio and pain measures. Both t-tests and Pearson correlations were carried out in R Software (R Core Team (2020). R: A language and environment for statistical computing. R Foundation for Statistical Computing, Vienna, Austria. URL https://www.R-project.org/) and figures were generated using the ggplot2 package [79].

## RESULTS

### Fibromyalgia Syndrome vs. Pain-Free Control Participants

E/I ratios in were significantly higher in FMS than in healthy controls, in relation to both water (Δ*M* = .084, *t*(18.307) = 2.219, *p* = .039) and Cr (Δ*M* = .085, *t*(18.303) = 2.241, *p* = .038; Figure 3). However, pain-free controls were significantly younger than people with FMS (Δ*M* = 10.4, *t*(65)= 2.703, *p* = .009). According to a study investigating GABA concentration differences in young vs. older adults, groupwise differences were attributable to bulk tissue composition within the MRS voxel and upon subsequent tissue volume correction were no longer statistically significant [48]. FMS (GM: *M* = 0.41, *SD* = 0.058; WM: *M* = 0.53, *SD* = 0.073) and control (GM: *M* = 0.40, *SD* = 0.061; WM: *M* = 0.56, *SD* = 0.079) cohorts did not demonstrate significant differences in either grey matter (GM; *t*(65) = −0.66, *p* = 0.51, *d* = .19) or white matter (WM; *t*(65) = 1.49, *p* = 0.15, *d* = .42) tissue composition within the MRS voxels (Figure 2). CSF fraction was not normally distributed, so a non-parametric Wilcoxon Rank Sum Test was used to assess group differences. Participants with FMS (*M* = 0.060, *SD* = 0.032) had a significantly greater fraction of CSF within the MRS voxels compared to pain-free controls (*M* = 0.040, *SD* = 0.026, *Z* = −2.4, *p* = 0.017, *d* = .71; Figure 2). In the female-only subsample, there were no differences in DLPFC GM or WM (*t*(57) = .026, *p* = .979 and *t*(57) = .885, *p* = .380), but CSF fraction was greater in FMS than pain-free controls (*t*(57) = 2.133, *p* = .037). To account for potential E/I ratio differences related to voxel tissue composition, segmentations (GannetSegment) were performed to correct for CSF content, GM and WM GABA content, and water relaxation as described by Harris and colleagues [29]. Using the fully corrected spectra in relation to water, E/I ratios were significantly higher in people with FMS than in healthy controls (*t*(18.305) = 2.223, *p* = .039). This difference remained significant in the female-only subsample (*t*(57) = 3.717, *p* < .001; Figure 4).

**Figure 3.**
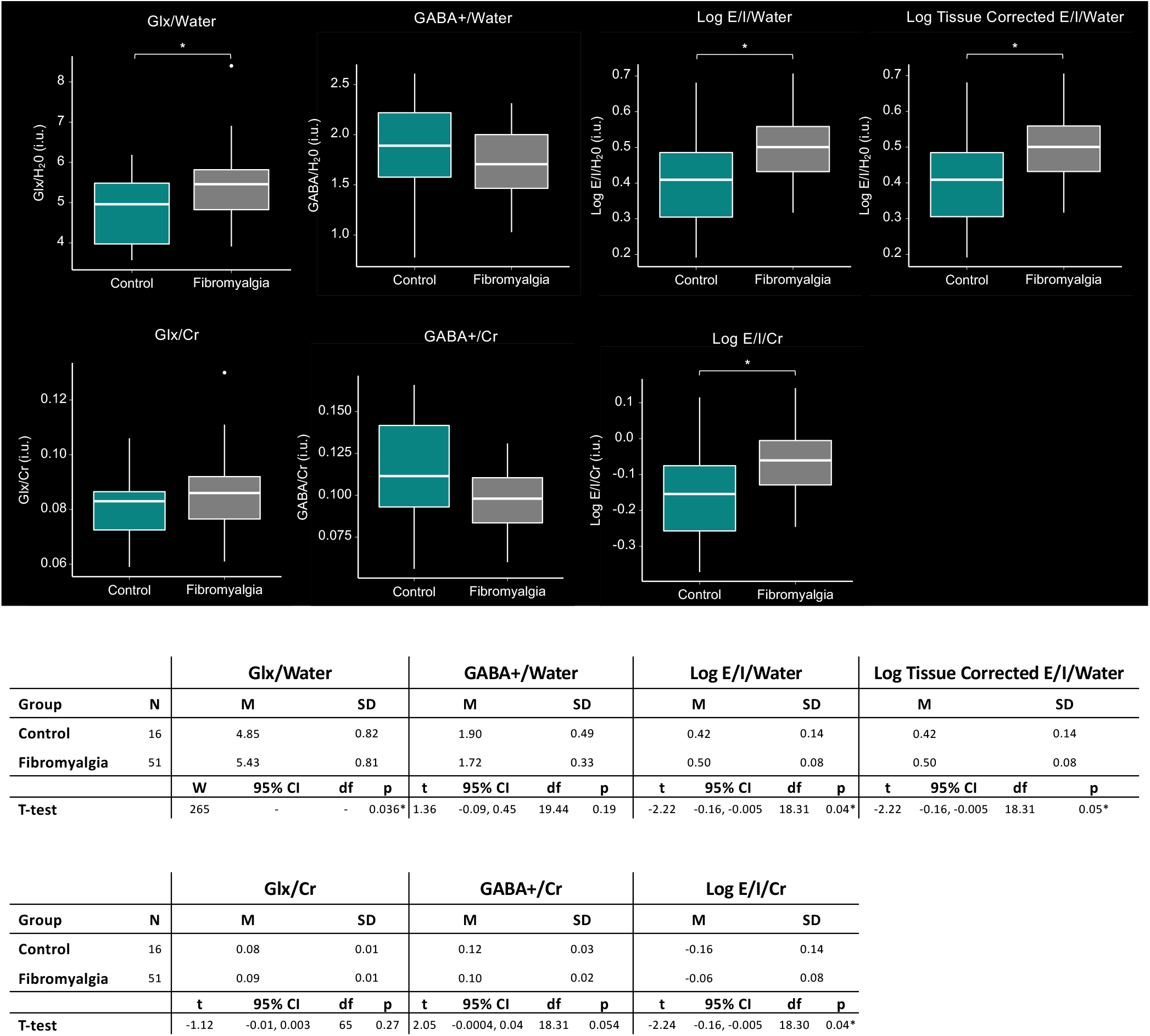
Between Group Comparison of L-DLPFC Neurochemical Concentration (Males and Females Combined): The boxplots and table illustrate comparisons in neurochemical concentrations between controls and participants with FMS relative to both creatine (Cr) and water in the combined male and female cohort. Across reference metabolites, there was a trend of increased Glx and decreased GABA+ in FMS which was combined to examine differences in the E/I ratio. E/I ratio was increased in individuals with FMS compared to controls across reference metabolites with and without correcting for tissue volume fractions.

**Figure 4.**
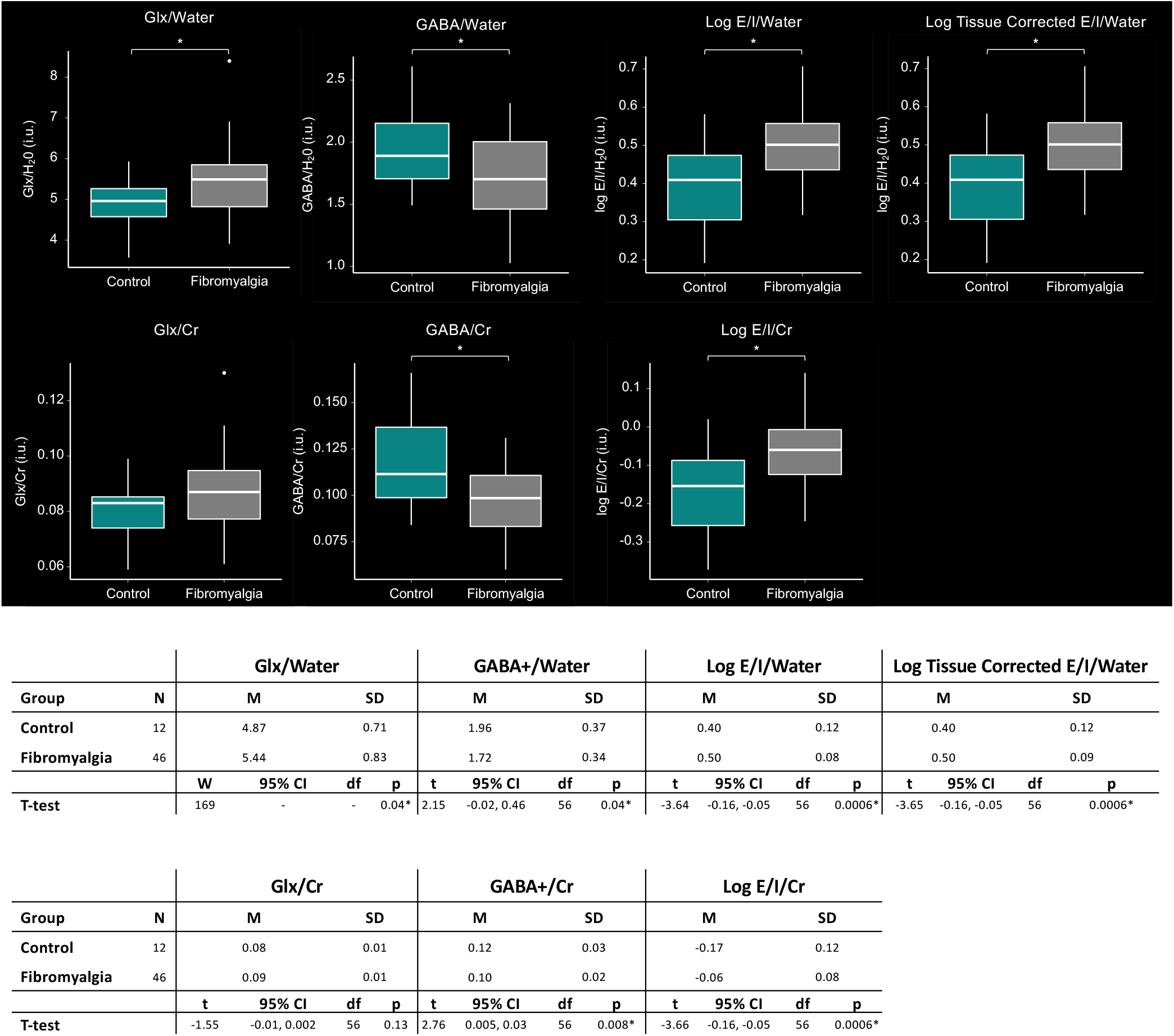
Between Group Comparison of L-DLPFC Neurochemical Concentration (Females Only): The boxplots and table illustrate comparisons in neurochemical concentrations between controls and participants with FMS relative to both creatine (Cr) and water the female only cohort. Across reference metabolites, there was a significant increase in Glx, decrease in GABA+, and increased E/I ratio with and without tissue volume correction in the FMS cohort.

Because depression has been shown to influence neurochemical concentrations, specifically within the DLPFC, an exploratory analysis was conducted in the FMS cohort by stratifying for subjective depression based on the Mini-Mental State Examination (MMSE) conducted as part of the screening procedures. It is important to note that the MMSE does not provide an indicator of depression severity, and suicidality was exclusionary. Nonetheless, consistent with the known prevalence of depression in FMS, 38 out of 51 participants were identified to have comorbid depression+FMS compared to 13 identified as FMS without depression. Controlling for age, the analysis of covariance (ANCOVA) demonstrated a significant effect of diagnosis (i.e. depression+FMS vs FMS without depression) on E/I ratios in Cr (*F*(1, 48) = 4.962, *p* = .031, *η*^*2*^ = .094), water (*F*(1, 48) = 5.084, *p* = .029, *η*^*2*^ = .096), and the corrected spectra in relation to water (*F*(1, 48) = 5.049, *p* = .029, *η*^*2*^ = .095), with comorbid depression+FMS being associated with lower E/I ratio than FMS alone. Age did not have a significant effect on any of the E/I ratios (Cr: *F*(1, 48) = .009, *p* = .924, *η*^*2*^ = .000; water and water-corrected: *F*(1, 48) = .010, *p* = .920, *η*^*2*^ = .000).

### E/I Ratios as Predictor of Pain Tolerance and Threshold

In participants with FMS, E/I ratios in relation to both water and Cr significantly predicted thermal pain tolerance respectively (*R*^*2*^ = .101, *F*(1,49) = 5.530, *β* = .318, *p* = .023 and *R*^*2*^ = .103, *F*(1,49) = 5.632, *β* = .321, *p* = .022) (Figure 5), in that greater E/I ratios were associated with greater pain tolerance. The water and Cr models indicate a non-significant trend (*p* = .071 and *p* = .068, respectively) when age was considered as a covariate. However, age was not a significant predictor (*p* = .697 and *p* = .698, respectively) and did not impact the relationship between thermal pain tolerance and E/I ratios in both water (*R*^*2*^ = .104, *F*(2,48) = 2.794, *β* = .320, *p* = .023) and Cr (*R*^*2*^ = .106, *F*(2,48) = 2.843, *β* = .322, *p* = .022).

**Figure 5.**
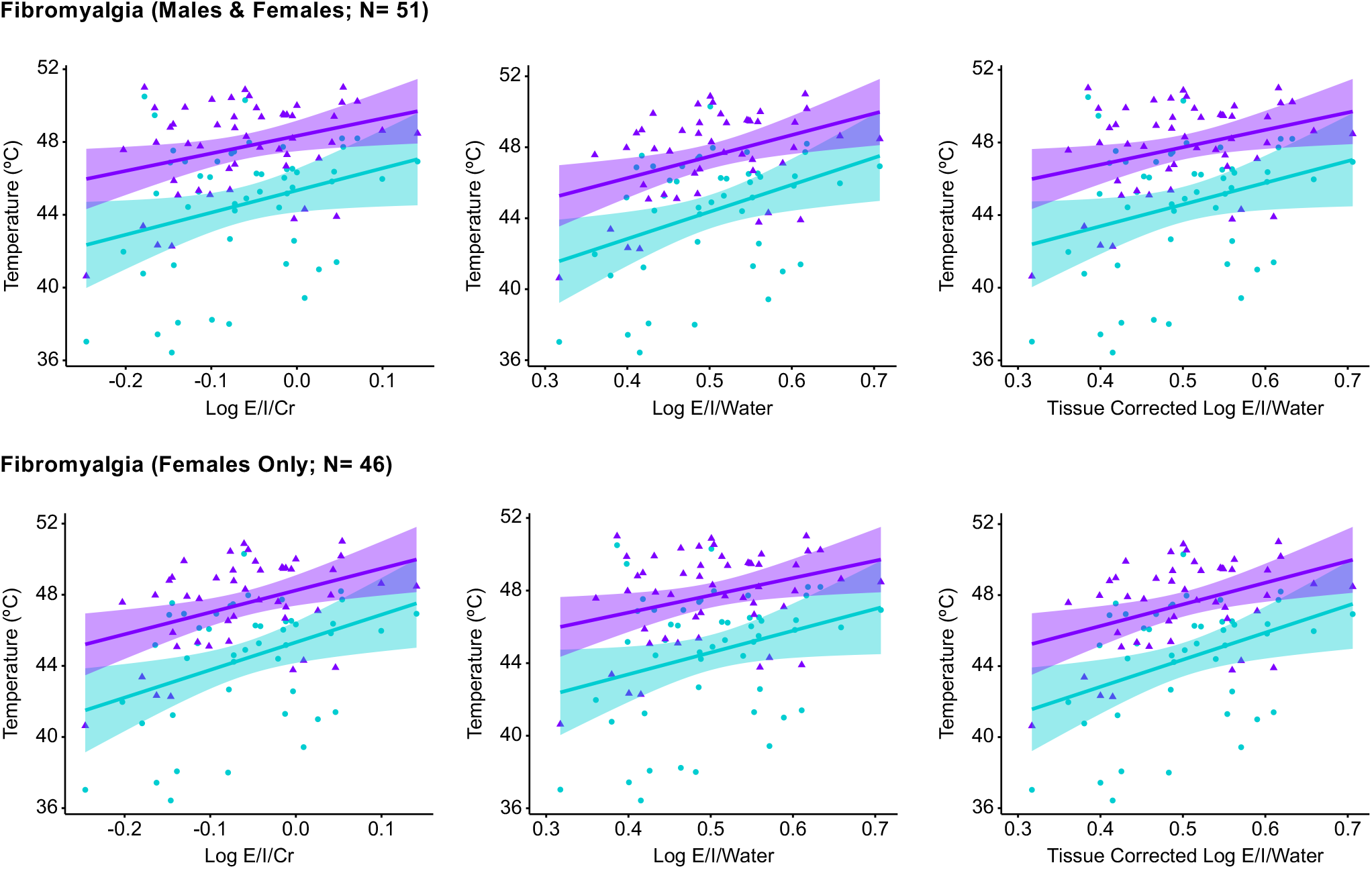
E/I Ratio as a Predictor of Pain Tolerance and Threshold in Participants with FMS: Regression scatterplots with 95% confidence intervals of acute thermal pain threshold (cyan) and tolerance (purple) as a function of L-DLPFC E/I ratio referenced to creatine and water with and without stratification by sex. An analysis of only male participants was not conducted due to the minimum sample size in the FMS cohort (n=5).

Similarly in direction to pain tolerance, the models significantly predict thermal pain threshold (water: *R*^*2*^ = .081, *F*(1,49) = 4.315, *β* = .284, *p* = .043; Cr: *R*^*2*^ = .083, *F*(1,49) = 4.425, *β* = .288, *p* = .041) (Figure 5), and indicated a non-significant trend (*p* = .071 and *p* = .090, respectively) when age was included as a covariate. Age was not a significant predictor (*p* = .417 in both models) and did not impact the relationship between thermal pain threshold and E/I ratios (water: *R*^*2*^ = .094, *F*(2,48) = 2.478, *β* = .287, *p* = .042; Cr: *R*^*2*^ = .095, *F*(2,48) = 2.532, *β* = .291, *p* = .040).

A subsequent analysis was conducted controlling for sex (i.e., looking at females only, as they comprised 46 out of 51 participants). In females with FMS, E/I ratios in relation to both water and Cr significantly predicted thermal pain tolerance (*R*^*2*^ = .158, *F*(1,44) = 8.270, *p* = .006 and *R*^*2*^ = .163, *F*(1,45) = 8.541, *p* = .005, respectively). The water and Cr models remained significant even after including age as a covariate (*R*^*2*^ = .159, *F*(2,43) = 4.050, *p* = .024 and *R*^*2*^ = .163, *F*(2,44) = 4.184, *p* = .022, respectively). E/I ratios were the only significant predictors in both the water (E/I: *β* = .398, *p* = .007; age: *β* = .017, *p* = .902) and Cr (E/I: *β* = .404, *p* = .006; age: *β* = .019, *p* = .895) models. There were no concerns of multicollinearity for any of the predictors (*Tolerance* = .998, *VIF* = 1.002), and the data met assumptions of independent errors (*Durbin-Watson* = 1.988). The models significantly predicted thermal pain threshold (water: *R*^*2*^ = .138, *F*(1,45) = 7.064, *β* = .372, *p* = .011; Cr: *R*^*2*^ = .143, *F*(1,45) = 7.342, *β* = .378, *p* = .010), and remained significant after considering age as a covariate (water: *R*^*2*^ = .140, *p* = .039; Cr: *R*^*2*^ = .145, *p* = .034). Age did not impact the relationship between thermal pain tolerance and E/I ratios in water (E/I: *β* = .374, *p* = .011; age: *β* = .046, *p* = .748) and Cr (E/I: *β* = .380, *p* = .010; age: *β* = .047, *p* = .741).

An additional post-hoc analyses was conducted to account for individual differences in pain experience (subjective pain), to contextualize the thermal pain tolerances. A composite score was created to include both objective (temperature tolerance) and subjective pain rating (average rating over five pain intervals: 1 = no pain, to 10 = worst possible pain). Scores were calculated as the mean Z score of both objective and subjective pain levels:

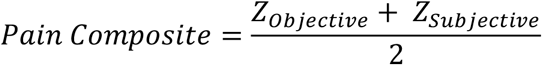

Pain composite scores in the whole sample were significantly predicted by E/I ratios in both water (*R*^*2*^ = .102, *F*(1,49) = 5.569, *β* = .319, *p* = .022) and Cr (*R*^*2*^ = .104, *F*(1,49) = 5.666, *β* = .322, *p* = .021). Both the water and Cr models showed a non-significant trend after considering age as a covariate (*p* = .071 and *p* = .068, respectively). As with the first analyses, including age as a covariate did not impact the relationship between thermal pain tolerance and E/I ratios in both water (*β* = .318, *p* = .024) and creatine (*β* = .321, *p* = .023).

### Association between E/I Ratio and Pain Metrics

Pearson correlation coefficients were computed to assess the relationship between E/I ratio and clinical pain symptoms assessed via the BPI short form collapsed across both FMS and control participants. In the combined male+female analyses, a significant positive correlation between BPI and E/I ratio in relation to Cr: *r(65) =*.*27, p <*.*029*, water: *r(65) =*.*26, p <*.*031*, and water with tissue correction: *r(65) =*.*26, p <*.*031* was identified (Figure 6). In the female only analyses, there was a significant positive correlation between BPI and E/I ratio in relation to Cr: *r(56) =*.*3, p <*.*021*, water: *r(56) =*.*3, p <*.*022*, and water with tissue correction: *r(56) =*.*3, p <*.*022*.

**Figure 6.**
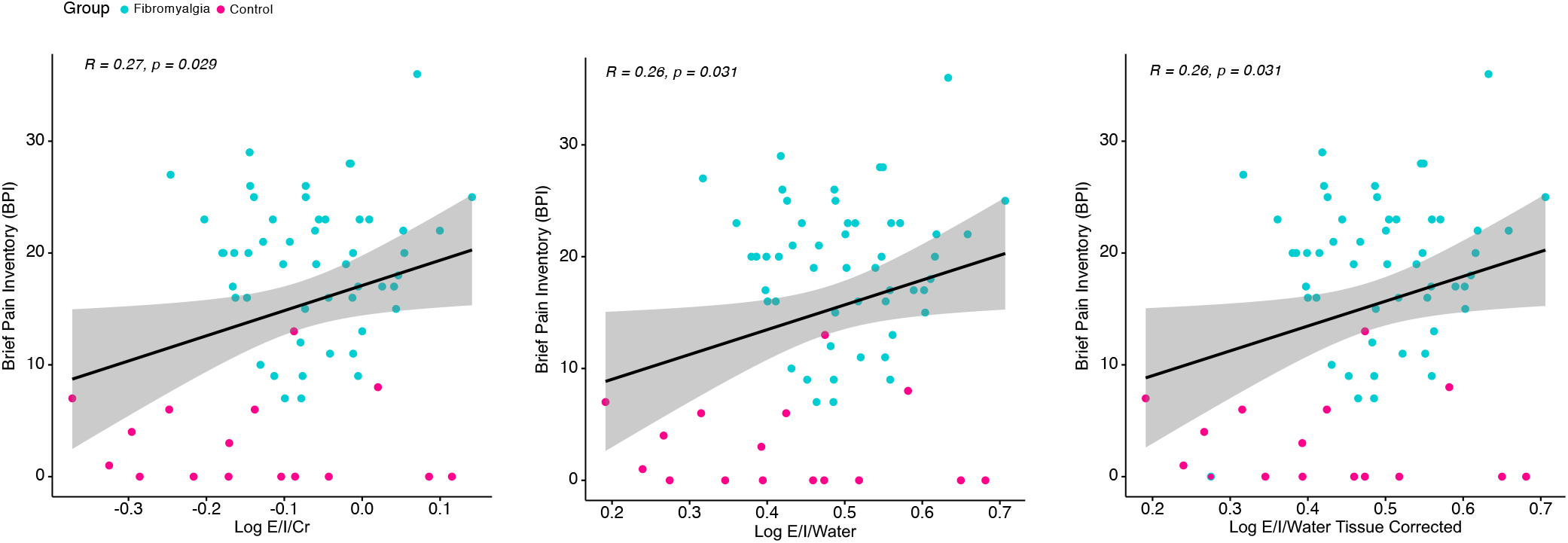
Relationship Between Clinical Pain and L-DLPFC E/I Ratio Across Pain and Controls: Scatterplots of clinical pain measured by the brief pain inventory as a function of E/I ratio collapsed across both FMS (cyan) and control (pink) cohorts.

## DISCUSSION

In this study we identified excitatory and inhibitory neurotransmitter imbalances within the L-DLPFC of participants with FMS compared to controls using ^1^H-MRS. This study expands upon previous findings of increased Glx and decreased GABA observed across pain-related regions by investigating neurochemical concentrations within the L-DLPFC. The findings in this report are novel and clinically relevant because they demonstrate combined dysfunction in the balance of excitation and inhibition within brain circuitry that could be targeted for potential interventions.

Both Glx and GABA have been implicated across chronic pain conditions - recently reviewed by Peek and colleagues [57]. The balance of excitation and inhibition serves a critical role in the pathogenesis of FMS by altering cortical excitability [51], mediating receptor up/downregulation, and contributing to pathological processes involved in promoting the maintenance of a pain state via central sensitization [41]. Centralized pain disorders such as FMS have previously been shown to exhibit alterations in both Glx and GABA respectively (see reviews [55; 57]). Because MRS investigations are often limited to single or several discrete regions of interest, as opposed to whole brain imaging common across functional and structural imaging modalities, the majority of studies to date in FMS have generally focused on primary sensory and affective pain circuitry including the thalamus [34], amygdalae [75], somatosensory, and insular cortices [31]. In these regions, elevated Glx has been consistently demonstrated across investigations in chronic pain cohorts. Within prefrontal cortical structures, reports of Glx alterations are varied. Specifically, Glx concentrations in the orbitofrontal cortex between FMS and pain-free controls did not differ [75]. Alternatively, our results are consistent with those of Feraco and colleagues who reported elevated Glx – albeit in the bilateral ventrolateral prefrontal cortices – relevant to executive control and emotion regulation [16].

There is currently little direct evidence in FMS demonstrating neurochemical dysfunction within the DLPFC. In other musculoskeletal pain disorders such as chronic low back pain, biochemical alterations in N-acetyl aspartate (NAA) and glucose have been identified within this structure [23; 24], posited to reflect neuronal density [21]. While NAA concentrations have been associated with pain measures in FMS and chronic fatigue syndrome they have not been shown to be significantly different from controls [61; 77]. Although glutamate excitotoxicity could result in alterations of NAA concentration [65], consistent with reduced grey matter volumes reported in FMS [38], recent multimodal imaging evidence does not corroborate compromised neuronal integrity [62] suggesting an alternative mechanism for increased glutamate concentrations.

Elevated excitatory tone induced artificially in rodents has been shown to manifest in increased pain sensitivity as well as reduced peripheral intraepidermal nerve fiber density [32]. This suggests a potential top-down mechanism for the disorder driven by increased CNS glutamate without inducing excitotoxicity. Our results indicate that a combination of metabolites representing the balance of both excitation and inhibition in FMS play a role in the pathophysiology of the condition both with and without comorbid depression. Current pharmacological strategies to treat FMS-related pain further support the role of glutamate/glutamine in the pathophysiology of the condition. For example, pregabalin has been shown to be an effective therapy for FMS in multiple clinical trials [12], and although the mechanisms of gabapentinoid drugs are complex and not fully characterized [73], treatment ameliorates aberrant Glx (glutamate/glutamine) levels measurable by MRS [30].

Our results support dysfunction in both neurotransmitter systems contributing to an altered ratio of excitation to inhibition in FMS. The conversion of glutamate to GABA is enzymatically mediated by glutamic acid decarboxylase (GAD), ultimately regulating GABA neurotransmission [52] and homeostasis [15]. GAD has two isoforms that include GAD65 (predominate isoform), important for regulating vesicular release of GABA, and GAD67 which functions as the primary metabolic driver of GABA production. GAD65 is associated with many of the symptoms and clinical characteristics common to individuals with FMS [17]. Knockout of GAD65 in mice confirms a supraspinal role of the enzyme in producing thermal hyperalgesia [37]. The prevalence of comorbid psychiatric conditions such as major depressive disorder is high in FMS and GAD function has also been implicated in neuropsychiatric conditions (see review: [35]) including reduced levels in the prefrontal cortex [59], although subsequent studies suggest that GAD concentrations in unipolar depression were similar to nonpsychiatric patient controls [25]. Nonetheless, animal models of mild chronic stress demonstrate reduced GAD activity and anhedonia-like behaviors [14] consistent with high co-morbidity of depression observed clinically in FMS. Studies investigating GAD levels in patients with FMS are limited to a single case report [74] and mention of co-occurrence in an examination of levels in a secondary condition [27]. Contributing factors of GAD dysregulation include both sleep disturbances and anxiety that frequently co-occur in FMS. Many of the collective hallmarks of FMS are supported by GAD dysregulation providing a potential mechanism of altered E/I concentrations that may provide a common link to the seemingly disparate cluster of idiopathic clinical features patients often present with. Future studies will be required to directly test the potential role that GAD plays on E/I dysfunction in FMS.

Glutamate and glutathione also contribute to the measured Glx peak. Treatment with the latter has been shown to attenuate mechanical allodynia in a rat model of centralized pain analogous to complex regional pain syndrome [83]. In patients with FMS, augmentation of glutathione, a prevalent antioxidant, with coenzyme Q10, reduced clinical pain and increased pressure pain threshold in addition to reducing neural activity [67]. With the current study methodology, it is not feasible to disentangle the specific contributions of Glx metabolites, however, elevated glutathione concentrations appear to be protective and related to diminished clinical pain suggesting the possible contribution of alternative molecular influences via glutamate and glutamine. Glutamine, a precursor to glutamate, is elevated in cerebrospinal fluid of individuals with FMS which was shown to be positively associated with evoked symptomatic tender points [40]. Glutamate is also elevated in the CSF of individuals with FMS [58] and is involved in enhanced excitability (central sensitization) and increased pain sensitivity [66]. Our findings exhibiting increased L-DLPFC Glx in FMS participants as well as the positive relationship between E/I ratio and clinical pain are in line with these previous findings.

Several reports in FMS populations have elucidated relationships between either Glx [31; 85] or GABA [18] individually. In the current experiment, the L-DLPFC subregion was defined via the strength of functional connectivity to the dACC, a hub of the salience network, and critical to pain processing [78]. Our results indicate a positive relationship between E/I ratio and acute pain threshold and tolerance, which has yet to be reported on in FMS but is counter to previous findings examining the relationship between Glx and pain sensitivity within the insula [31]. ACC E/I ratio has been shown to be positively associated with depression and anxiety symptoms in veterans with pain [43] – both of which are frequently reported in FMS. Nonetheless, a potential explanation for discrepancy in relationship includes the spatial location of MRS measurements which could influence neurochemical concentration [6], however, the right DLPFC was included in pooled findings demonstrating a positive association between Glx and pain sensitivity in healthy (pain-free) individuals [85]. Alternatively, in an earlier finding, Glx was negatively associated with pressure threshold [31], although the relationship was examined by collapsing across pain and control groups. Discrepant alterations in metabolite concentrations have been reported during acute compared to tonic pain in healthy participants using functional MRS (MRS acquired during a task) [10; 39]. Additionally, investigations in FMS are frequently conducted in limited FMS populations (i.e., female participants only). Sex differences in neuroimaging findings have been shown across chronic pain conditions including FMS [26]. Although the study sample included both male and female participants, exploratory analyses of the female cohort alone revealed consistent relationships between E/I ratio. Insufficient sample size precluded comparison of sex differences between males and females, and thus subsequent studies will need to be conducted to investigate potential sex-specific neurochemical alterations in FMS.

In addition to neural excitability, alterations in glutamatergic transmission are implicated in glia reactivity with astrocytes playing a pivotal role in the tripartite synapse which regulates glutamate-GABA turnover (see review; [28]). Increased glial reactivity in chronic pain conditions, including FMS, has been identified in structures including the DLPFC using PET imaging as well as MRS approaches capable of resolving myoinositol, a marker of glial cell proliferation [1; 7; 46] - with higher levels associated with pain-evoked functional connectivity [36]. Specifically, Pomares and colleagues demonstrated increased [18F]flumazenil PET ligand uptake, which binds to GABAA receptors prevalent at cortical inhibitory synapses [63]. From these studies it is not feasible to determine whether receptor-level or metabolic alterations precede one another, however, GABAA receptor upregulation could manifest from diminished GABA concentrations or vice versa. Our results demonstrating an increase in Glx, decrease in GABA, and increased E/I ratio (Glx/GABA) are in line with the possibility of dysregulation in glutamate-GABA turnover (discussed above) with both imbalances of excitatory/inhibitory neurotransmission and/or glia involvement as potential mechanisms. These theories are speculative given the current data, and further investigations are required to provide a more definitive mechanistic link between E/I dysfunction and FMS.

### Caveats

#### Overlap with co-morbid disorders

FMS is a heterogeneous disorder with a spectrum of underlying symptoms and high comorbidities with depression, anxiety/stress disorders, as well as cognitive and motor deficits. Independent of FMS, these conditions could influence Glx and GABA concentrations particularly within cortical regions such as the DLPFC. GABA is reduced in the prefrontal cortex in both preclinical models of depression and clinical populations with major depressive disorder [19]. This is relevant to chronic pain, given the high comorbidity of depression (lifetime prevalence up to 90%) and anxiety among of patients with FMS [22; 49; 56]. This co-incidence is increased as pain severity worsens [54]. Glutamate dysregulation has been widely reported in MDD, underscored by the glutamate hypothesis for depression, which is consistent with effective therapies such as ketamine, that in part, act as an NMDA receptor antagonist [53]. To examine whether depression influenced Glx and GABA, we conducted an exploratory post-hoc analysis categorizing the FMS cohort using the MMSE by: 1) FMS with depression and 2) FMS without depression. Due to both the categorical and subjective definition of depression obtained using the MMSE, cautious interpretation of effects is warranted. An effect of diagnosis was identified suggesting that the influence of comorbid psychiatric disorders on E/I ratio cannot be ruled out. Future studies, using continuous scales to elucidate severity of depression or other co-morbid disorders are necessitated to determine the influence and/or relationship on E/I ratio in the L-DLPFC.

## CONCLUSIONS

We provide evidence of combined excitatory/inhibitory imbalance within the L-DLPFC in FMS, which was demonstrated to be positively associated with acute/clinical pain measures. Together these results suggest a dysregulation of excitation and inhibition in top-down pain modulatory networks with potential pathophysiological implications in pain processing.

## Data Availability

All data produced in the present study are available upon reasonable request to the authors.

## ACKNOWLEDGEMENTS & DISCLOSURES

This work was supported by the NIH National Center for Complementary and Integrative Health grants 5R33AT009305-03 (NW/DS) and 1F32AT010420-01 (JB). The Stanford University Molecular Imaging Scholars Fellowship (T32CA118681; JB) and the Center for Neurobiological Imaging Innovation Award (JB). We would like to thank Katy Stimpson, Merve Gulser, Catherine Tran, Ruben Krueger, and Anthony Toves for help with participant recruitment, MRI data collection, and/or data management. The authors would also like to thank Keith Sudheimer, Ph.D. for fMRI technical assistance.

Dr. Williams is a named inventor on Stanford-owned intellectual property relating to neuroimaging-based TMS targeting; he has served on scientific advisory boards for Otsuka, NeuraWell, Nooma, and Halo. The other authors declare no competing financial or conflicts of interests.

---

**Supplementary Figure 1.**
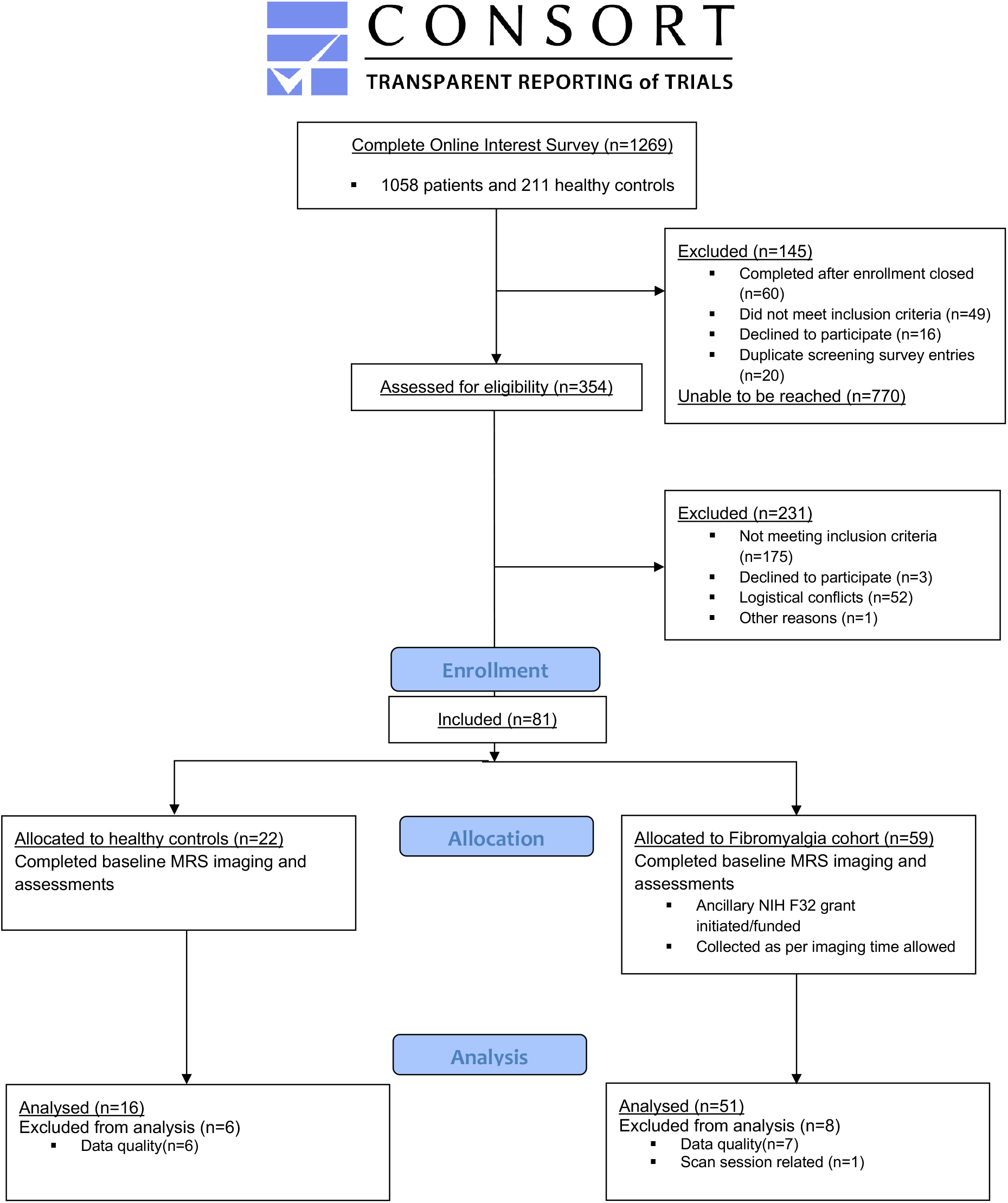
Consort Diagram.

